# Empathy and clarity in GPT-4-Generated Emergency Department Discharge Letters

**DOI:** 10.1101/2024.10.07.24315034

**Authors:** Gal Ben Haim, Adva Livne, Uri Manor, David Hochstein, Mor Saban, Orly Blaier, Yael Abramov Iram, Moran Gigi Balzam, Ariel Lutenberg, Rowand Eyade, Roula Qassem, Dan Trabelsi, Yarden Dahari, Ben Zion Eisenmann, Yelena Shechtman, Girish N Nadkarni, Benjamin S Glicksberg, Eyal Zimlichman, Anat Perry, Eyal Klang

## Abstract

**Background and Aim:** The potential of large language models (LLMs) like GPT-4 to generate clear and empathetic medical documentation is becoming increasingly relevant. This study evaluates these constructs in discharge letters generated by GPT-4 compared to those written by emergency department (ED) physicians.

**Methods:** In this retrospective, blinded study, 72 discharge letters written by ED physicians were compared to GPT-4-generated versions, which were based on the physicians’ follow-up notes in the electronic medical record (EMR). Seventeen evaluators, 7 physicians, 5 nurses, and 5 patients, were asked to select their preferred letter (human or LLM) for each patient and rate empathy, clarity, and overall quality using a 5-point Likert scale (1 = Poor, 5 = Excellent). A secondary analysis by 3 ED attending physicians assessed the medical accuracy of both sets of letters.

**Results:** Across the 72 comparisons, evaluators preferred GPT-4-generated letters in 1,009 out of 1,206 evaluations (83.7%). GPT-4 letters were rated significantly higher for empathy, clarity, and overall quality (p < 0.001). Additionally, GPT-4-generated letters demonstrated superior medical accuracy, with a median score of 5.0 compared to 4.0 for physician-written letters (p = 0.025).

**Conclusion:** GPT-4 shows strong potential in generating ED discharge letters that are empathetic and clear, preferable by healthcare professionals and patients, offering a promising tool to reduce the workload of ED physicians. However, further research is necessary to explore patient perceptions and best practices for leveraging the advantages of AI together with physicians in clinical practice.

## INTRODUCTION

The growing integration of large language models (LLMs) such as GPT-4 into healthcare is reshaping medical practice (1). GPT-4 has demonstrated capabilities in clinical reasoning and performed comparably to physicians on standardized exams like the United States Medical Licensing Examination (USMLE) (2,3). Notably, GPT-4 has also performed similarly to humans on USMLE empathy questions (4). Furthermore, GPT-4 achieves similar medical scores to emergency department (ED) physicians, underscoring its potential in real-world clinical environments (5,6).

EDs worldwide are facing increasing pressure due to growing patient volumes and limited amounts of clinical practitioners, driven by an aging population and the escalating complexity of medical care (7). Physicians frequently face significant time constraints, exacerbated by the demands of administrative tasks, such as writing detailed discharge letters, which prolongs patient throughout. Automating aspects of this documentation process could reduce these burdens, allowing ED physicians to devote more time to direct patient care (8–10).

Empathy - broadly defined as understanding the other, sharing their emotional experience and caring for them (11,12) is crucial in the physician–patient relationship and has a well-documented impact on healthcare outcomes. Greater physician empathy is associated with improved clinical outcomes, higher patient satisfaction, and better adherence to treatment (13,14). For example, empathy has been shown to predict patient cooperation and compliance with treatment plans, emphasizing its importance in medical practice (15). Hojat et al. even demonstrated that empathy correlates with improved outcomes for diabetic patients (16).

Recent studies suggest that GPT-4 can generate responses that are perceived as empathetic when interacting with patients, raising important questions about its potential to replicate human-like communication. While previous studies have compared physician responses to AI-generated ones in hypothetical scenarios, online experiments, or online advice forums, this study takes an ecological approach by comparing physician and AI-generated responses in a real, high-stakes clinical setting—the ED. Moreover, while previous studies used either physicians or the general population as raters (17), we focused on three highly relevant groups who need to understand these discharge letters: the ED physicians who work together, the nurses who often communicate the results to the patients, and the patients themselves, who not only take these recommendations home but are expected to adhere to the prescribed treatment and follow-up care. (11,16,).

To this end, this study compared the empathy and clarity conveyed in discharge letters generated by GPT-4 with those written by ED physicians. By focusing on empathy in a real-world clinical setting, we seek to determine whether AI can both enhance the clarity as well as the emotional quality of medical documentation in emergency care. Understanding this capability is pivotal for considering implementation into clinical workflows.

### Study Design

#### Study Design

This was a retrospective, blinded, comparative study designed to evaluate the empathy, clarity, and quality of discharge letters generated by GPT-4 compared to those written by human physicians. The study was conducted on April 3, 2024, in the Emergency Department (ED) of Sheba Medical Center, a large tertiary care hospital in Israel. The study also included a secondary evaluation to ascertain the medical accuracy of GPT-4-generated letters.

#### Population and Data Collection

The study included all patients discharged from the ED during an 8-hour nurse shift (8 a.m. to 3 p.m.). For each patient, two discharge letters were created: one written by the attending ED physician and another generated by GPT-4 based on patients’ medical records, which serve for clinical purposes and are therefore considered highly reliable. All data were retrieved after an institutional review board (IRB) permission was granted.

#### Artificial Intelligence Framework

A standardized prompt was used to generate each GPT-4 letter, simulating the perspective of a senior ED physician. The prompt provided the LLM with de-identified follow-up notes, originally written in Hebrew, from the ED visit within the EMR, excluding the discharge letter, and instructed it to produce a concise medical summary and discharge recommendations, if applicable. The web-based version of GPT-4 was used, with a new instance generated for each prompt in May 2024. The following template was applied for all cases:

I am conducting a survey comparing the empathy of physicians with that of GPT-4. You are a senior physician in the emergency department. I will provide you with the medical information, and you will have decided to discharge the patient. Please write a concise medical summary in free text (avoid bullet points), including specific discharge recommendations, if necessary. Be professional and empathetic, ensuring that empathy does not compromise professionalism. Please write in English and then translate into Hebrew.

#### Evaluator Recruitment and Blinding

A total of 17 evaluators were recruited for the study, including 7 physicians (4 residents and 3 specialists), 5 patients, and 5 nurses, Evaluators were drawn from a mix of staff within the institute and external individuals, including random ED patients. To minimize bias, evaluators were blinded both to the source of the discharge letters and to the purpose of the study. They were presented with side-by-side comparisons of the original human-written letter (L1) and the GPT-4-generated letter (L2), with the order of the letters randomized, without any indication that one letter was AI-generated.

#### Evaluation Process

The evaluation process was divided into two components:

1. Primary Evaluation (Empathy, Clarity and Quality Assessment): Evaluators were asked to compare the two letters for each patient using a structured questionnaire.
First, evaluators made a binary choice indicating which letter they preferred overall (L1 or L2). Additionally, the following criteria were assessed on a 5-point Likert scale for each letter (1 = Poor, 5 = Excellent):
  - Empathy: The extent to which the letter conveyed compassion and understanding.
  - Clarity of Summary: How clear and understandable the medical summary was.
  - Overall Quality: The overall quality of the letter, reflecting its effectiveness in communication.
  - Clarity of Recommendations: How clear and actionable the discharge instructions were.
2. Secondary Evaluation (Medical Accuracy): Three different senior ED attendings independently assessed the medical accuracy of both physician-written and GPT-4- generated discharge letters. The evaluators were blinded to the source of the letters, unaware of which were AI-generated. Accuracy was rated on a 5-point scale, based on how closely each letter aligned with the original ED notes for the patient.

#### Outcome Measures

Primary Outcomes: The primary outcomes were the clarity, quality and perceived empathy of GPT-4-generated discharge letters compared to those written by ED physicians.

Secondary Outcomes: separate analyses for each group (physicians, nurses, patients). As well medical accuracy of the letters, as rated by independent evaluators.

#### Statistical Analysis

Ordinal variables (e.g., empathy, clarity, and quality scores) were summarized using medians and interquartile ranges (IQR). Categorical variables (e.g., binary preference for L1 or L2) were reported as proportions. Statistical comparisons for ordinal data were made using the Mann-Whitney U test, while binary comparisons were made using chi-square tests. In addition, we performed a qualitative analysis of empathy in the humans and AI letters. All analyses were performed using Python. Statistical significance was defined as p < 0.05.

## RESULTS

### Study Characteristics

During a single emergency department shift, 72 patients were discharged, each receiving a discharge letter that formed the basis of our study. These letters, crafted by human professionals were compared to GPT-4 crafted letters. A diverse group of 17 evaluators, comprising physicians, patients and nurses, participated in the assessment (**Table 1**). The evaluators represented a wide range of experience and backgrounds, with an age distribution ranging from 25 to 74 years and the majority (53%) in the 35–44 age group. This varied group allowed for a broad perspective on the evaluation of empathy, clarity, and quality in the discharge letters.

**Table 1:** Study evaluators’ characteristics.

| Gender | Age | Classification | Specialty |
| --- | --- | --- | --- |
| Female | 35-44 | Physician,<br>Resident | Emergency<br>Medicine |
| Male | 35-44 | Physician,<br>Resident | Internal<br>Medicine |
| Female | 25-34 | Physician,<br>Resident | Emergency<br>Medicine |
| Male | 25-34 | Physician,<br>Resident | Emergency<br>Medicine |
| Female | 25-34 | Physician,<br>Resident | Emergency<br>Medicine |
| Female | 35-44 | Physician,<br>Expert | Emergency<br>Medicine |
| Female | 35-44 | Physician,<br>Expert | Oncology |
| Female | 25-34 | Nurse | Emergency<br>Medicine |
| Male | 35-44 | Nurse | Emergency<br>Medicine |
| Male | 35-44 | Nurse | Emergency<br>Medicine |
| Male | 25-34 | Nurse | Emergency<br>Medicine |
| Female | 45-54 | Nurse | Emergency<br>Medicine |
| Female | 35-44 | Patient | - |
| Male | 65-74 | Patient | - |
| Male | 45-54 | Patient | - |
| Male | 45-54 | Patient | - |
| Female | 35-44 | Patient | - |

### Overall Preference for Letters (GPT-4 vs Human)

Across 72 comparisons between the GPT-4-generated letter (L2) and the human-written letter (L1), evaluators expressed a strong preference for L2. Out of the total 1,206 recorded comparisons, L2 was preferred in 1,009 instances (83.7%), while L1 was preferred in 197 instances (16.3%). This substantial difference in preference suggests a marked favorability towards the GPT-4-generated discharge letters (p < 0.001). When conducting the same analysis for each evaluator-group separately (**Figure 1**), we found that GPT-4 was also consistently preferred across all groups (all p<0.001).

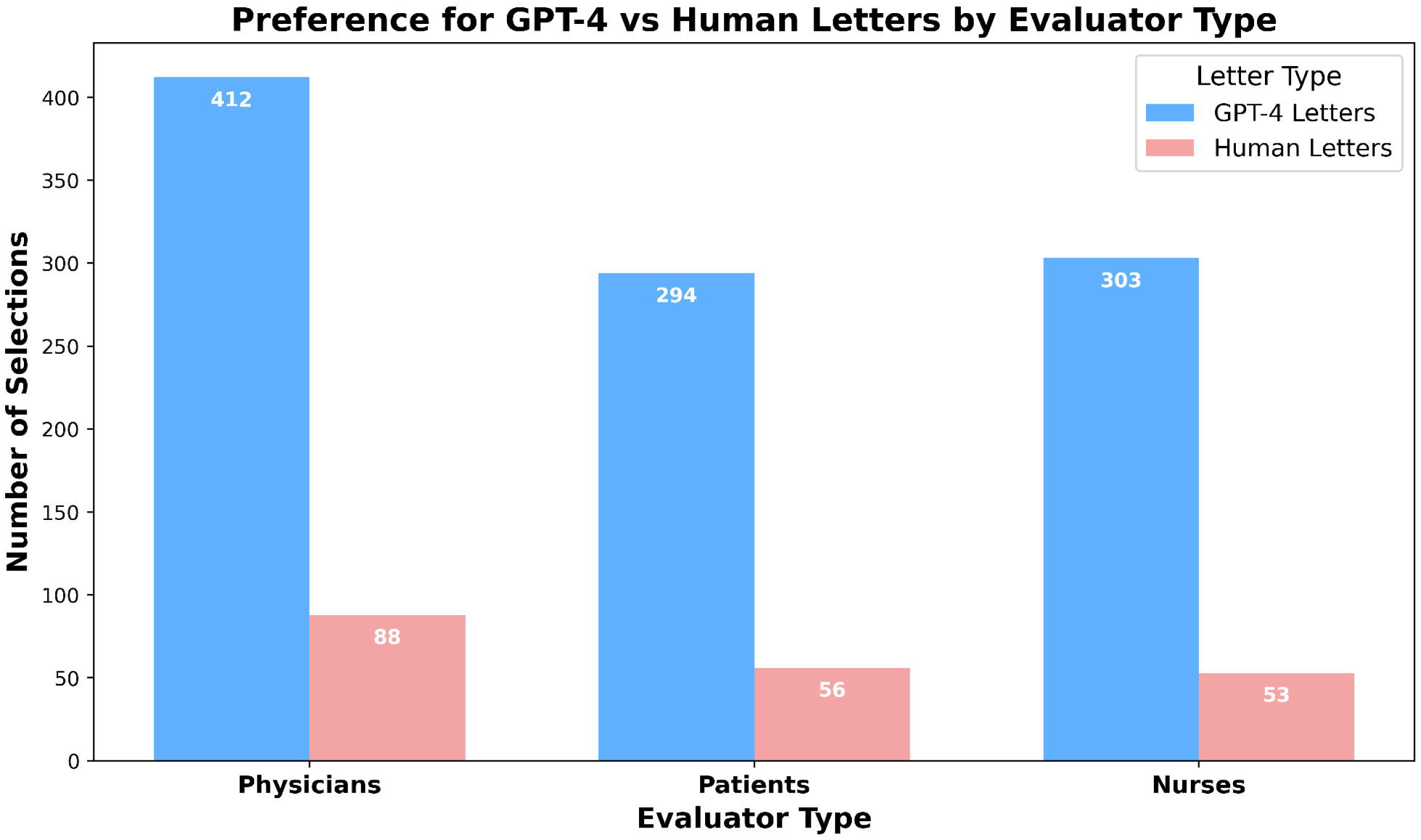

### Comparison of Ratings for GPT-4 and Human-Written Discharge Letters

Across all evaluator groups, GPT-4-generated discharge letters were rated significantly higher than human-written letters in most categories (**Table 2**). Statistically significant differences were observed in ratings for empathy, quality, and clarity of the summary (all p < 0.001). The only exception was for clarity of recommendations in the patients’ group, where no significant difference was found (p = 0.771). In the overall analysis, GPT-4 letters consistently outperformed human-written letters across all metrics (p < 0.001), reinforcing the trend seen in individual evaluator groups.

**Table 2:** Median (IQR) scores of Empathy, Quality, and Clarity for human letters vs GPT-4 letters by evaluator group.

| Evaluator Group | Empathy |  | Quality |  | Clarity of Summary |  | Clarity of Recommendations |  |
| --- | --- | --- | --- | --- | --- | --- | --- | --- |
|  | Human | GPT-4 | Human | GPT-4 | Human | GPT-4 | Human | GPT-4 |
| Physicians | 3.0 (2.5 - 3.5) | 4.0 (4.0 - 4.5) | 3.0 (3.0 - 3.5) | 4.0 (4.0 - 4.5) | 3.0 (3.0 - 4.0) | 4.0 (4.0 - 4.5) | 4.0 (3.0 - 4.0) | 4.0 (3.5 - 4.5) |
| Patients | 3.0 (2.0 - 3.0) | 4.0 (3.0 - 4.0) | 3.0 (3.0 - 3.0) | 4.0 (4.0 - 4.0) | 3.0 (3.0 - 4.0) | 4.0 (4.0 - 4.0) | 4.0 (4.0 - 4.0) | 4.0 (3.0 - 4.0) |
| Nurses | 2.0 (2.0 - 3.0) | 4.0 (3.0 - 4.0) | 3.0 (2.0 - 3.0) | 4.0 (3.0 - 4.0) | 3.0 (2.0 - 3.0) | 4.0 (3.0 - 4.0) | 3.0 (3.0 - 4.0) | 4.0 (3.5 - 5.0) |
| Overall | 3.0 (2.0 - 3.0) | 4.0 (3.0 - 4.0) | 3.0 (2.0 - 4.0) | 4.0 (4.0 - 5.0) | 3.0 (2.0 - 4.0) | 4.0 (3.0 - 4.0) | 4.0 (3.0 - 4.0) | 4.0 (3.0 - 5.0) |

### Subgroup Analysis Based on Gender and Age

Overall, GPT-4 letters consistently received higher ratings across most subgroups and metrics, with a few exceptions in the clarity of recommendations for certain groups (**Table 3**).

**Table 3:**
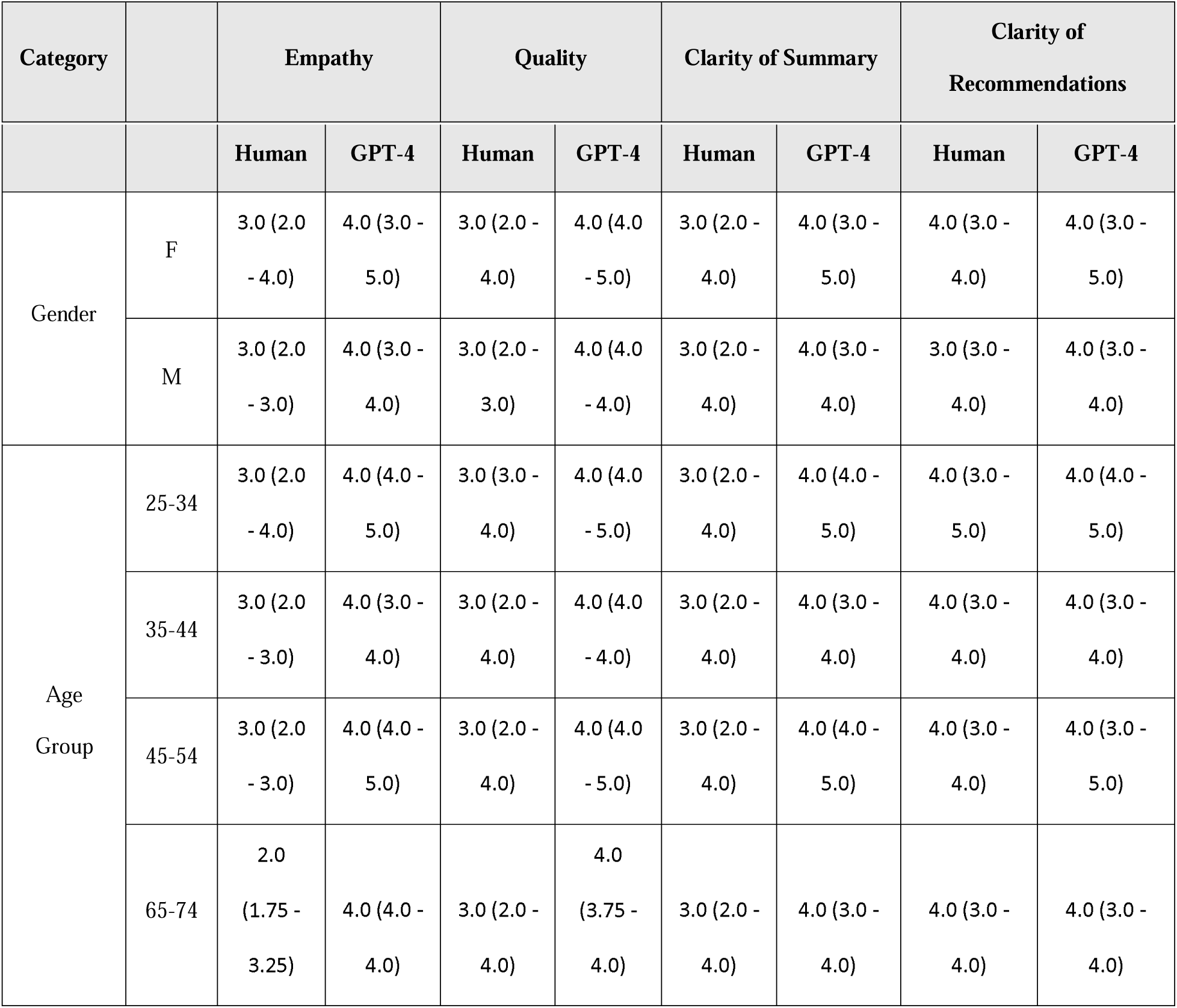
Median (IQR) scores of Empathy, Quality, and Clarity for human letters vs GPT-4 letters by evaluator categories.

For **gender**, both females and males rated GPT-4 letters significantly higher than human-written letters across all metrics. Females showed a particularly strong preference for GPT-4 in empathy, quality, and clarity of the summary (p < 0.001), with a smaller but still significant difference in clarity of recommendations (p < 0.001). Males also rated GPT-4 significantly higher across empathy, quality, and clarity (p < 0.001). Given evidence suggesting that women are more attuned to empathy, these traits may hold greater importance for female evaluators, potentially explaining their heightened sensitivity to empathetic elements in the discharge letters (19).

When broken down by **age**, GPT-4 letters were consistently rated higher for empathy, quality, and clarity in the 35-44, 25-34, and 45-54 age groups (p < 0.001 for most metrics). However, in the 65-74 age group, while GPT-4 letters were rated significantly higher for empathy, quality, and clarity of the summary (p < 0.001), no significant difference was found for clarity of recommendations (p = 0.723). Similarly, for the 45-54 age group, no significant difference was found in clarity of recommendations (p = 0.265), despite higher ratings for other metrics.

### Accuracy Evaluation

In the evaluation of the accuracy of ED discharge letters, the three ED attendings rated both human-written (L1) and GPT-4-generated (L2) letters on a scale of 1-5. The results, summarized in **Table 4**, show that the overall median accuracy score for L1 was 4.0 (IQR 4.0 - 4.5), while L2 received an overall higher median score of 5.0 (IQR 4.0 - 5.0). This finding was statistically significant with p = 0.025, indicating that GPT-4 letters were at least as accurate, if not better, compared to human-written letters.

**Table 4:**
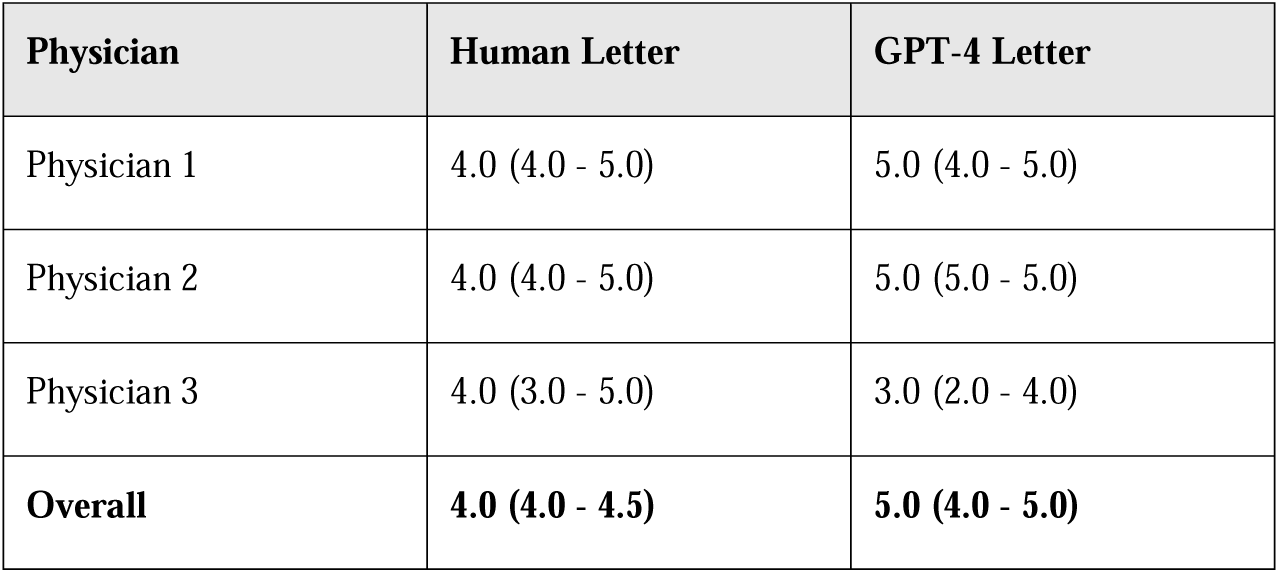
Median (IQR) scores of accuracies for human letters vs GPT-4 letters by ED attendings evaluator categories.

### Qualitative Analysis

A qualitative comparison of GPT-4-generated and physician-written discharge letters revealed differences in how empathy was conveyed.

In one physician-written letter for a patient with a chronic condition, the physician expressed personalized support: “I understand that managing your symptoms has been challenging, but we are here to support you every step of the way. Please contact us if you experience any new symptoms.” In contrast, the GPT-4-generated letter stated: “Your condition has been stable, and we recommend follow-up care with your primary physician if there are any changes.” While accurate, the GPT-4 letter lacked the personal reassurance seen in the physician’s note.

However, in other cases, GPT-4 demonstrated a sufficient level of empathy. For example, a GPT-4 letter for a patient with a recent cancer diagnosis included: “We know this diagnosis may be overwhelming, and we are committed to helping you through this difficult time. Please reach out with any concerns.” While this language may be viewed as more general compared to the physician’s note for the same case, which was more clinical and concise, evaluators still rated the GPT-4 letter higher for its clarity and perceived empathy.

**Table 5** provides 15 examples of how empathy was conveyed in both AI and physician-written letters. Although GPT-4’s responses may sometimes seem to rely on standardized expressions of empathy, evaluators consistently favored these letters over those written by physicians. In these cases, GPT-4’s approach, though may be described as “generic” or “standardized,” was not seen as a limitation by the majority of evaluators and was overwhelmingly preferred overall (L1 vs. L2).

**Table 5:** This table captures the contrast in communication styles and levels of empathy between the AI-generated and physician-written discharge letters.

| Example # | Case Description | GPT-4 | Physician (Human) |
| --- | --- | --- | --- |
| 1 | Chronic condition | "Managing your symptoms is challenging, but we are here to support you." | "Your condition has been stable. Follow up if changes occur." |
| 2 | Cancer diagnosis | "We know this diagnosis may be overwhelming. We are committed to helping you." | "Your tests indicate a cancer diagnosis. Follow up with oncology." |
| 3 | Respiratory distress | "Your symptoms have improved. Continue your treatment plan." | "Patient stable. Follow instructions given upon discharge." |
| 4 | Surgery follow-up | "Your post-surgery recovery is on track. Follow up in two weeks." | "Post-surgery status stable, follow up in clinic." |
| 5 | Pain management | "We understand that chronic pain is difficult, and we're here to support you." | "Pain management stable, continue medications as prescribed." |
| 6 | Diabetes management | "It's important to manage your glucose carefully, and we know it can be tough. Reach out if you need help." | "Glucose levels normal, continue regular monitoring." |
| 7 | New diagnosis | "We understand that receiving a new diagnosis can be overwhelming. We're here to support you." | "New diagnosis explained, follow up for further care." |
| 8 | Hypertension | "Managing blood pressure can be challenging, but you're doing a great job. Keep it up." | "BP well controlled, continue medication as prescribed." |
| 9 | Stroke recovery | "We understand that stroke recovery is difficult, and we're here to help." | "Patient recovering well post-stroke. Continue therapy." |
| 10 | Heart failure | "Living with heart failure can be overwhelming. Reach out if you have concerns." | "Condition stable, follow up with cardiologist." |
| 11 | Mental health crisis | "We know this has been hard, and we're here to support you." | "Patient stable, continue psychiatric follow-up." |
| 12 | Migraine treatment | "We understand that migraines can affect your daily life. We're here to help you manage them better." | "Patient stable, continue current treatment." |
| 13 | Asthma exacerbation | "Asthma can be difficult to manage. We're committed to helping you breathe easier." | "Symptoms resolved, continue medication as needed." |
| 14 | Post-ER discharge | "We hope you're feeling better, and remember, we're here if you need anything." | "Discharged in stable condition. Follow-up as instructed." |
| 15 | Chronic pain | "We recognize the difficulty of living with chronic pain and are committed to finding a solution." | "Pain under control, continue medications." |

## Discussion

This study highlights the potential of GPT-4 in generating high-quality, empathetic and clear discharge letters within the emergency department (ED) setting. Evaluators consistently preferred GPT-4-generated letters over physician-written ones, particularly in terms of empathy, clarity, and overall quality. Both physicians and non-physician evaluators rated GPT-4 higher across most metrics, with empathy and clarity emerging as key strengths.

Previous studies have demonstrated that large language models (LLMs), such as GPT variants, can surpass medical experts in summarizing clinical texts and generating discharge summaries (9,20,21). These models produce content comparable to human-written summaries, with patients often perceiving AI-generated instructions as equally clear, albeit occasionally lacking the personal touch found in human-written documents (18). In this study, GPT-4-generated letters were also rated more accurate than those written by physicians. This suggests that AI tools like GPT-4 may enhance the accuracy of medical documentation, which is crucial for patient safety and outcomes.

Our study is unique in its focus on full discharge letter generation, in a real high-stakes clinical setting, with a particular emphasis not just on accuracy but mainly on empathy and clarity — key aspects of effective patient communication. By assessing these qualities in a real-world ED setting, we offer insight into AI’s ability to balance both technical accuracy and emotional connection in clinical documentation.

Interestingly, there were no significant differences between groups when it came to the clarity of the discharge recommendations. This is a positive finding for physicians, suggesting that while the summaries they write may be less empathetic or clear in other aspects, their discharge recommendations remain consistently clear across all groups.

The high empathy ratings for GPT-4 are consistent with prior research showing that AI can generate responses perceived as compassionate, even in clinical settings (12,17). This is especially relevant in high-stress environments like the ED, where empathy plays a crucial role in guiding patients through the next phase of care, including self-care and follow-up with community physicians. The sense of being seen and cared for may significantly enhance patient adherence to treatment and follow-up care (22,23).

Although GPT-4’s approach may be viewed as “generic” in some cases, this did not detract from its effectiveness. On the contrary, evaluators overwhelmingly favored the AI-generated letters, suggesting that GPT-4’s standardized yet empathetic responses were seen as more helpful and communicative. In a direct, blinded comparison, GPT-4 consistently outperformed human-written letters across multiple metrics. This underscores a simple reality: when tested head-to-head, GPT-4 not only held its own against physicians but often surpassed them, demonstrating that even “standardized” empathy can be more impactful in practice.

One caveat to consider is that empathy ratings were assessed without disclosing the identity of the empathizer. Some studies have indicated that when AI authorship is disclosed, the perceived empathy of the response diminishes (18). This could be attributed to bias against AI (24) but also to a fundamental human need for care from fellow humans—those capable of potentially sharing in their pain or sorrow and genuinely caring for them (25). This underscores the need for caution when implementing AI in direct patient care, even if it performs on par or better than humans in certain areas (18,24). However, AI should be seen as a tool to assist physicians with administrative tasks, with the physician maintaining final control over the content, editing, and signing of the discharge letter.

Thus, by generating discharge letters, GPT-4 shows potential to reduce the administrative burden on ED physicians by automating this time-consuming task. This could allow physicians to spend *more time* on direct patient care, which is critical in increasingly overcrowded and complex ED environments (26,27). Together, these results highlight the importance of striking the right balance between leveraging the strengths of AI and preserving the unique qualities of human connection and care—whether by using AI as a co-pilot or ensuring a human-in-the-loop, (though even this may come with a cost) (18).

Despite the promising findings, there are limitations to this study. The evaluators were from a single demographic, and the study was conducted at a single center, limiting the generalizability of the results. Moreover, we did not employ advanced techniques such as retrieval-augmented generation or fine-tuning, which could have further improved GPT-4’s performance. Despite this, GPT-4’s zero-shot performance was rated highly by the evaluators, indicating strong baseline capabilities. Additionally, while GPT-4 performed well in this study, further research is needed to understand how patients perceive AI-generated discharge letters, when they know that the letters were generated by AI, or by AI and physicians together. Lastly, future studies should explore whether cultural or institutional differences influence the reception of AI-generated content.

In conclusion, GPT-4 shows strong potential in generating ED discharge letters that are empathetic and clear, preferable by healthcare professionals and patients, offering a promising tool to reduce the workload of ED physicians. However, further research is necessary to explore patient perceptions and best practices for leveraging the advantages of AI together with physicians in clinical practice.

## Data Availability

All data produced in the present study are available upon reasonable request to the authors.

## References

1. Dave T, Athaluri SA, Singh S. ChatGPT in medicine: an overview of its applications, advantages, limitations, future prospects, and ethical considerations. Front Artif Intell. 2023;6:1169595.

2. Gilson A, Safranek CW, Huang T, Socrates V, Chi L, Taylor RA, et al. How Does ChatGPT Perform on the United States Medical Licensing Examination (USMLE)? The Implications of Large Language Models for Medical Education and Knowledge Assessment. JMIR Med Educ. 2023 Feb 8;9:e45312.

3. Kung TH, Cheatham M, Medenilla A, Sillos C, De Leon L, Elepaño C, et al. Performance of ChatGPT on USMLE: Potential for AI-assisted medical education using large language models. PLOS Digit Health. 2023 Feb;2(2):e0000198.

4. Brin D, Sorin V, Vaid A, Soroush A, Glicksberg BS, Charney AW, et al. Comparing ChatGPT and GPT-4 performance in USMLE soft skill assessments. Sci Rep. 2023 Oct 1;13:16492.

5. Haim GB, Braun A, Eden H, Burshtein L, Barash Y, Irony A, et al. AI in the ED: Assessing the efficacy of GPT models vs. physicians in medical score calculation. Am J Emerg Med. 2024 May;79:161–6.

6. Lahat A, Sharif K, Zoabi N, Patt YS, Sharif Y, Fisher L, et al. Assessing Generative Pretrained Transformers (GPT) in Clinical Decision-Making: Comparative Analysis of GPT-3.5 and GPT-4. J Med Internet Res. 2024 Jun 27;26(1):e54571.

7. Kenny JF, Chang BC, Hemmert KC. Factors Affecting Emergency Department Crowding. Emerg Med Clin North Am. 2020 Aug;38(3):573–87.

8. Maleki Varnosfaderani S, Forouzanfar M. The Role of AI in Hospitals and Clinics: Transforming Healthcare in the 21st Century. Bioeng Basel Switz. 2024 Mar 29;11(4):337.

9. Barash Y, Klang E, Konen E, Sorin V. ChatGPT-4 Assistance in Optimizing Emergency Department Radiology Referrals and Imaging Selection. J Am Coll Radiol JACR. 2023 Oct;20(10):998–1003.

10. Ben-Haim G, Yosef M, Rowand E, Ben-Yosef J, Berman A, Sina S, et al. Combination of machine learning algorithms with natural language processing may increase the probability of bacteremia detection in the emergency department: A retrospective, big-data analysis of 94,482 patients. Digit Health. 2024;10:20552076241277673.

11. Zaki J, Ochsner KN. The neuroscience of empathy: progress, pitfalls and promise. Nat Neurosci. 2012 Apr 15;15(5):675–80.

12. Sorin V, Brin D, Barash Y, Konen E, Charney A, Nadkarni G, et al. Large Language Models (LLMs) and Empathy – A Systematic Review [Internet]. medRxiv; 2023 [cited 2024 Oct 3]. p. 2023.08.07.23293769. Available from: https://www.medrxiv.org/content/10.1101/2023.08.07.23293769v1

13. Decety J. Empathy in Medicine: What It Is, and How Much We Really Need It. Am J Med. 2020 May;133(5):561–6.

14. Kim SS, Kaplowitz S, Johnston MV. The effects of physician empathy on patient satisfaction and compliance. Eval Health Prof. 2004 Sep;27(3):237–51.

15. Rakel DP, Hoeft TJ, Barrett BP, Chewning BA, Craig BM, Niu M. Practitioner empathy and the duration of the common cold. Fam Med. 2009;41(7):494–501.

16. Hojat M, Louis DZ, Markham FW, Wender R, Rabinowitz C, Gonnella JS. Physicians’ empathy and clinical outcomes for diabetic patients. Acad Med J Assoc Am Med Coll. 2011 Mar;86(3):359–64.

17. Ayers JW, Poliak A, Dredze M, Leas EC, Zhu Z, Kelley JB, et al. Comparing Physician and Artificial Intelligence Chatbot Responses to Patient Questions Posted to a Public Social Media Forum. JAMA Intern Med. 2023 Jun 1;183(6):589–96.

18. Reis M, Reis F, Kunde W. Influence of believed AI involvement on the perception of digital medical advice. Nat Med. 2024 Jul 25;

19. Eisenberg N, Lennon R. Sex differences in empathy and related capacities. Psychol Bull. 1983;94(1):100–31.

20. Van Veen D, Van Uden C, Blankemeier L, Delbrouck JB, Aali A, Bluethgen C, et al. Adapted large language models can outperform medical experts in clinical text summarization. Nat Med. 2024 Apr;30(4):1134–42.

21. Falis M, Gema AP, Dong H, Daines L, Basetti S, Holder M, et al. Can GPT-3.5 generate and code discharge summaries? J Am Med Inform Assoc JAMIA. 2024 Oct 1;31(10):2284–93.

22. Moudatsou M, Stavropoulou A, Philalithis A, Koukouli S. The Role of Empathy in Health and Social Care Professionals. Healthcare. 2020 Jan 30;8(1):26.

23. Mercer S, Reynolds W. Empathy and quality care. Vol. 52 Suppl, The British journal of general practice : the journal of the Royal College of General Practitioners. 2002. S9 p.

24. Ragot M, Martin N, Cojean S. AI-generated vs. Human Artworks. A Perception Bias Towards Artificial Intelligence? In 2020.

25. Perry A. AI will never convey the essence of human empathy. Nat Hum Behav. 2023 Nov;7(11):1808–9.

26. Sorin V, Barash Y, Konen E, Klang E. Large language models for oncological applications. J Cancer Res Clin Oncol. 2023 Sep;149(11):9505–8.

27. Khoury A. Burnout syndrome in emergency medicine: it’s time to take action. Eur J Emerg Med Off J Eur Soc Emerg Med. 2022 Aug 1;29(4):239–40.

